# Culture-independent identification and serotyping of *Streptococcus pneumoniae* by targeted metagenomics in pleural fluid samples

**DOI:** 10.64898/2026.04.13.26350812

**Authors:** Sarah A.M. Smith, Rebecca J Rockett, Shahin Oftadeh, Kingsley King-Gee Tam, Michael Payne, Tanya Golubchik, Vitali Sintchenko

## Abstract

2.

*Streptococcus pneumoniae* is the leading cause of empyema and pneumonia in children, and monitoring of effectiveness of polyvalent pneumococcal vaccines has been essential for controlling invasive pneumococcal disease (IPD) in children and elderly adults. Conventional serotyping of pneumococci has relied on Quellung reaction following laboratory culture, however more recently whole genome sequencing (WGS) has been implemented in many reference laboratories to enhance traditional typing. Pleural fluid samples from cases with empyema are often culture negative, limiting the utility of WGS and requiring polymerase chain reaction (PCR) or 16S rRNA sequencing to detect *S. pneumoniae*. These molecular methods have limited sensitivity and capacity to characterise pneumococcus in clinical samples, especially in specimens with a low pathogen abundance. This study applied capture-based enrichment (tNGS) to identify and characterise *S. pneumoniae* directly from pleural fluid samples. A total of 51 pleural fluid samples were subjected to tNGS with a custom probe panel, for 39 known positive fluids collected from IPD cases between 2018-2025 in New South Wales, Australia. tNGS results were benchmarked against molecular-based serotyping. Our tNGS achieved 100% sensitivity and specificity in detecting *S. pneumoniae*. Serotyping results were concordant with PCR and 95% (37/39) of *S. pneumoniae* PCR positive pleural fluid cases could be serotyped using tNGS. Standard molecular methods however could only determine serotype in 56% (22/39) of samples. This tNGS enabled 39% improvement in ability to directly identify and serotype IPD-associated serotypes of *S. pneumoniae* in difficult-to-culture pleural fluids can significantly enhance laboratory surveillance of IPD as well as our understanding of vaccine effectiveness.

**Impact statement:** There is currently a gap in understanding the pneumococcus serotype diversity causing infection within the pleural fluid space. The gold-standard Quellung method to determine serotype relies on culturing the pneumococci first. However, pleural fluids often remain culture-negative, and cases of pneumococcal empyema have been a historical ‘blind spot’ in pneumococcal surveillance. This study offers a new methodology to close this gap and allow serotyping of previously untypable cases. The study demonstrated a targeted next generation sequencing (tNGS) approach to determine serotype without the need to first culture the bacteria. This novel use of tNGS targets part of the *cps* gene cluster, which determines serotype. To the best of our knowledge this is the first panel to do so. We have successfully serotyped 95% of pleural fluid *S.pneumoniae* PCR positive samples, where previously only 56% could be determined using conventional PCR typing methods. This demonstrates for the first time a novel tNGS method capable of determining the full serotype landscape causing pleural fluid infection. This development will enhance the understanding of vaccine effectiveness and contribute to the prevention of invasive pneumococcal disease.

**Data summary:** Supplementary data containing reference *cpsB* genomes are available within this article.

The authors confirm all supporting data, code and protocols have been provided within the article or through supplementary data files.

**Repositories:** ENA project accession number PRJEB111154. All supporting data has been provided within the article or in supplementary data files. One supplementary data file is available with the online version of this article.

## 5. Introduction

*Streptococcus pneumoniae* is responsible for invasive pneumococcal disease (IPD) which remains the leading cause of empyema and pneumonia in children, especially in those under five years of age (1, 2). Empyema occurs when the pneumococci invade the pleural fluid space. Cases of complicated pneumonia and empyema have been increasing internationally (3) and became a major cause of morbidity and mortality in developed countries (4). There are over 100 different serotypes of *S.pneumoniae* (5), but the current polyvalent conjugate vaccines (PCV13, PCV15 & PCV20), used in Australia (6) and internationally (7-9), cover only up to 20 of the most invasive serotypes. Since the introduction of pneumococcal vaccines there has been an increase in non-vaccine serotypes (10), however their association with empyema and distribution varies by geography (3).

Pleural fluid samples have been one of the most challenging samples for successful bacterial culture (11). This is partly due to prompt antibiotic therapy given to patients with empyema prior to fluid collection (12, 13), as these infections are often severe or even life-threatening (14). Such treatment reduces bacterial abundance in pleural fluid samples, making *S.pneumoniae* laboratory culture and subsequent serotyping difficult. In such circumstances, molecular methods such as polymerase chain reaction (PCR)-based serotyping (15) and 16S rRNA sequencing are often deployed to detect IPD and causitive serotypes (16, 17). However, these methods may not accurately detect *S.pneumoniae* serotype due to a low abundance of target DNA in pleural fluids, and biases in serotype-specific molecular methods (18, 19).

Here we validate a novel capture-based targeted metagenomics (tNGS) methodology which enables identification and characterisation of *S.pneumoniae* to the serotype level directly from pleural fluid samples, including cases where conventional culture is not informative. tNGS enriches pathogen-specific genomic regions through hybridisation with complementary biotinylated oligonucleotide probes, enabling the selective capture of low-abundance sequences. This approach allows sensitive detection and characterisation of pathogens using substantially fewer sequencing reads than required for shotgun metagenomics. This method has a higher sensitivity compared to traditional metagenomics (20, 21) and improves understanding of epidemiology of difficult-to-culture pathogens (22). In the past, similar targeted metagenomics approaches have been employed to identify specific genes of interest including those responsible for antimicrobial resistance (23). tNGS however, has yet to be utilised to determine serotype within clinical samples, which is essential to improve completeness of IPD surveillance.

We expanded a previously validated multi-pathogen probe capture panel, Castanet, which included probes targeting the ribosomal multi-locus sequence typing (rMLST) genes of *S.pneumoniae* (24), to include additional probes to target specific diagnostic and serotyping markers of *S.pneumoniae*. Since rMLST alone is insufficient to determine serotype (25), and may not be sufficiently specific in the presence of mixed respiratory flora, the expanded panel, Castanet 2.0, additionally included specific diagnostic loci (pneumolysin, *ply*, autolysin, *lytA*, and a putative transcriptional regulator gene,*SP2020*). These loci have been previously validated as targets for diagnostic PCR assays for *S.pneumoniae* detection (26). To determine serotype, we also included probes for the *cpsB* locus. The *cpsB* gene is part of the *cps* gene cluster (27) and the gold-standard locus used for molecular sequetyping assays (28). Following deep sequencing, reads corresponding to the *cpsB* target were used to determine serotype using kallisto (29), which performs reference-based competitive mapping of sequenced reads, assigning each read to its best-matching sequetype reference. We evaluated and compared this novel tNGS method with conventional molecular methods that have been used to identify serotypes within pleural fluid samples.

## 6. Methods

### 6.1 Validation sample cohort

*S.pneumoniae-*specific PCR positive, culture negative pleural fluid samples (n=44) collected from IPD cases between 2018-2025 were included in the study. These were collected and Quellung-based serotyping was performed at the New South Wales (NSW) Pneumococcal Reference Laboratory (PRL), Institute of Clinical Pathology and Medical Research (ICPMR), NSW Health Pathology.Pneumococcal serotypes were determined using 16S rRNA sequencing or serotype-specific PCR (29). In addition, pleural fluid samples that were culture negative for bacterial organisms (n=5) or positive for other bacteria capable to cause pleural fluid infections (n=6, *Listeria monocytogenes, Staphylococcus aureus, Streptococcus vestibularis, Streptococcus intermedius and Corynebacterium striatum*) were also sequenced as negative controls and to determine specificity of the tNGS method.

Four pure cultures of *S.pneumoniae* of serotypes 9N, 11A, 19F, and 19A, serotyped using the Quellung method, were used to assess the ability of the tNGS to detect and differentiate mixed serotype infections. Briefly, nucleic acid extracts from these four cultures were mixed in known proportions by volume, equally to achieve 25% abundance of each serotype or two cultures were mixed to simulate 12%, 25%, 50%, 75% and 88% abundance of each serotype.

### 6.2 Assessment of empyema burden in NSW Australia

The National Notifiable Diseases Surveillance System (NNDSS) Public Pneumococcal Dataset (2009–2024) was used to determine the abundance and serotype distribution of pleural infections within NSW. IPD cases recorded as either pleural effusion or empyema were used to examine serotype dynamics and age distribution.

### 6.3 Nucleic acid extraction

All pleural fluid clinical samples were extracted using Zymo Quick DNA/RNA high throughput kit (Zymo, California, USA) as per the manufacturer’s instructions. Following clinical specimen extraction, a diagnostic PCR was performed to determine quantity and presence of *S.pneumoniae* prior to tNGS library preparation and sequencing. Genomic DNA was extracted from pure *S.pneumoniae* cultures using the Ultraclean DNeasy microbial kit (Qiagen, Aarhus, Denmark), as per manufacturer’s instructions. Total DNA concentrations were quantified using Qubit™ 1X dsDNA HS Assay (Invitrogen™, ThermoFisher Scientific, Massachusetts, USA) allowing serotype proportions to be calculated.

### 6.4 Library preparation, capture and sequencing

All libraries were prepared using the Library Preparation Enzymatic Fragmentation (EF) kit v.1.0 (Twist Biosciences, South San Francisco, USA) and indexed using the TruSeq compatible Twist UMI Adapter System (Twist Biosciences, South San Francisco, USA). This was followed by a 17-hour capture, or hybridisation, using Twist Target Enrichment Standard Hybridisation and the Castanet 2.0 probe panel (24). Library preparation quantities were determined using Qubit™ 1X dsDNA HS Assay (Invitrogen™, ThermoFisher Scientific, Massachusetts, USA) and fragment sizes were ascertained by Agilent HS D5000 ScreenTape (Agilent Technologies Inc., California, USA). Samples were sequenced using a P1 flow cell on the NextSeq 2000 instrument (Illumina).

### 6.5 Bioinformatic analysis

All sequencing statistics including deduplicated reads counts, target coverage and depth were determined via the Castanet pipeline (v9.2) (30). This pipeline removes all human reads and summarises coverage on probe targets for each of the 35 bacteria enriched for in the panel. Pseudoalignment of reads to determine pneumococcal serotype abundance was performed using Kallisto (v0.48.0) (29). A reference index was created with a comprehensive collection of *cpsB* sequetyping reference sequences, on average 732-bp in length, for each serotype (n=396) (31). In addition, the *cpsB* sequetyping region of *Streptococcus mitis, Streptococcus oralis*, and *Streptococcus australis*, was also included, as these share genomic similarity with the *cpsB* region of *S.pneumoniae* (Supplementary data). Transcripts per million (TPM) measure was used to quantify the relative abundance of serotypes within pleural fluids as it normalises the abundance of reads mapping to targets within the reference index. All TPM values for each reference within a sample add to 1 million, allowing abundance of serotypes to be determined. In addition, mapped reads from Kallisto pseudobam output were analysed with Samtools (v1.9) (29) to determine the coverage and mean depth of reads mapping to the sequetyping regions within the reference index. A minimum coverage of 80% and read depth of 5x were required for each specimen to distinguish all informative single nucleotide polymorphisms (SNPs) for tNGS sequetyping analysis.

### 6.6 Descriptive statistics

Statistical analysis including correlations, t-tests, interquatile range (IQR) and median statistics, and result visualisation was conducted in R (v.4.5.1) using packages ggplot2 (v.4.0.0), ggmosaic (v.0.3.3) dplyr (v.1.1.4), tidyr (v.1.31), rstatix (v.0.7.2).

## 7. Results

### 7.1 Burden of pneumococcal empyema in Australia

A total of 402 cases of pleural empyema’s were recorded between 2009-2024 in New South Wales, Australia, accounting for 4.5% of the total IPD infections. Children under the age of 5 made up the largest age group experiencing pleural empyema at 55% (220/402). The most common serotype causing pleural infections was identified as *S.pneumoniae* serotype 3, at 42% (93/220) of all other clinical presentations in children under the age of 5, in NSW (220/1209) (Figure 1). Of these, 35% (77/220) of pleural empyema’s could not be serotyped. Comparatively, only 10% (97/989) of IPD cases with any other clinical presentation could not be serotyped.

**Figure 1.**
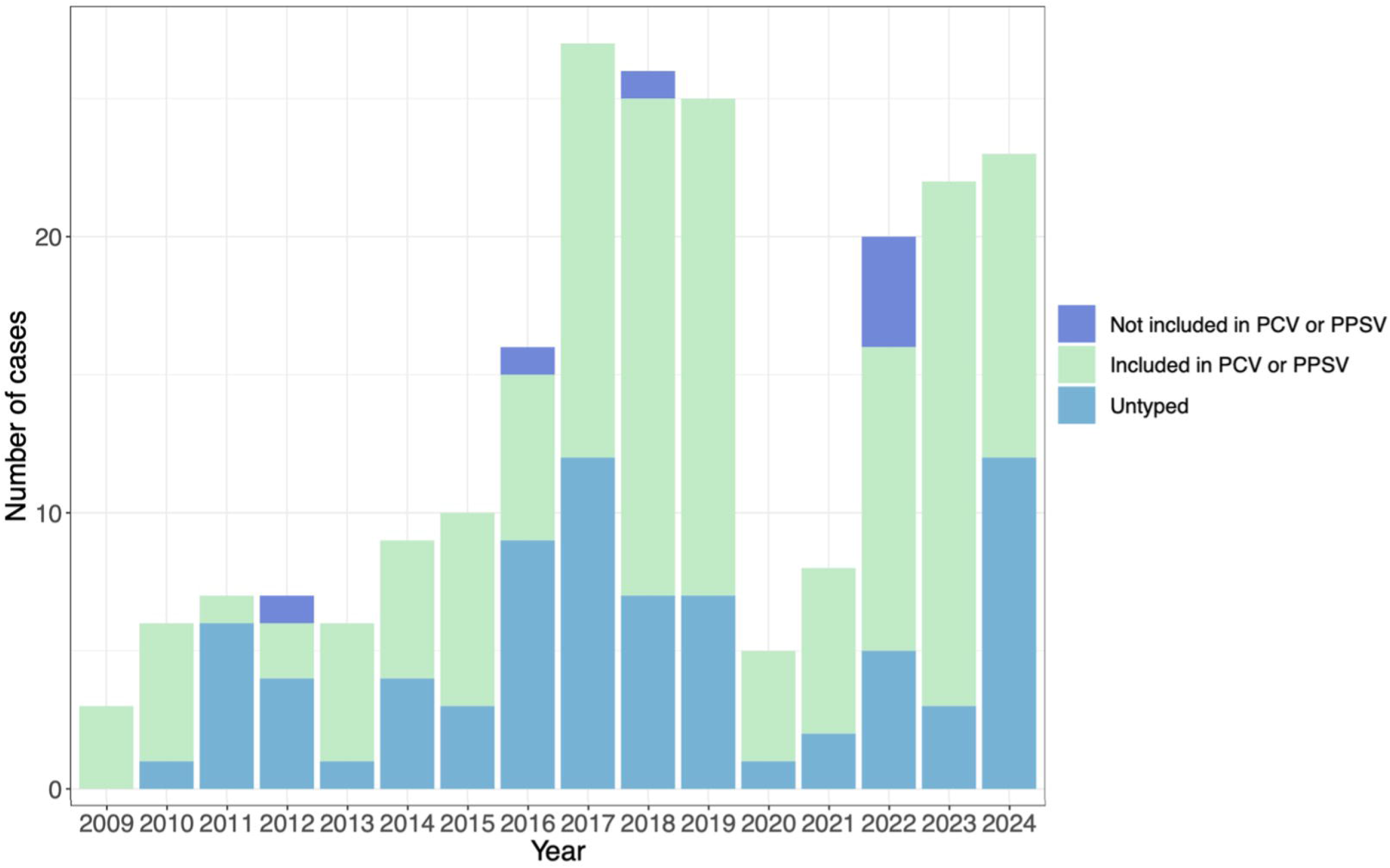
The frequency of vaccine and non-vaccine serotypes recovered from pleural effusion or empyema in children under the age of 5 in NSW, using molecular serotyping methods.

### 7.2 Detection of mixed pneumococcal populations by tNGS

Pneumococcal cultures of serotypes 9N, 11A, 19F, and 19A were mixed in pre-determined proportions and underwent sequencing and capture with the Castanet probe panel. They represented serotypes included in vaccine formulations and not-vaccine serotypes. All individual serotypes within the same sample were correctly identified (Figure 2A). Observed abundance closely matched expected (R = 0.96, Spearman correlation, p < 0.001), demonstrating that composition of serotypes within each sample could be successfully distinguished by tNGS (Figure 2B).

**Figure 2.**
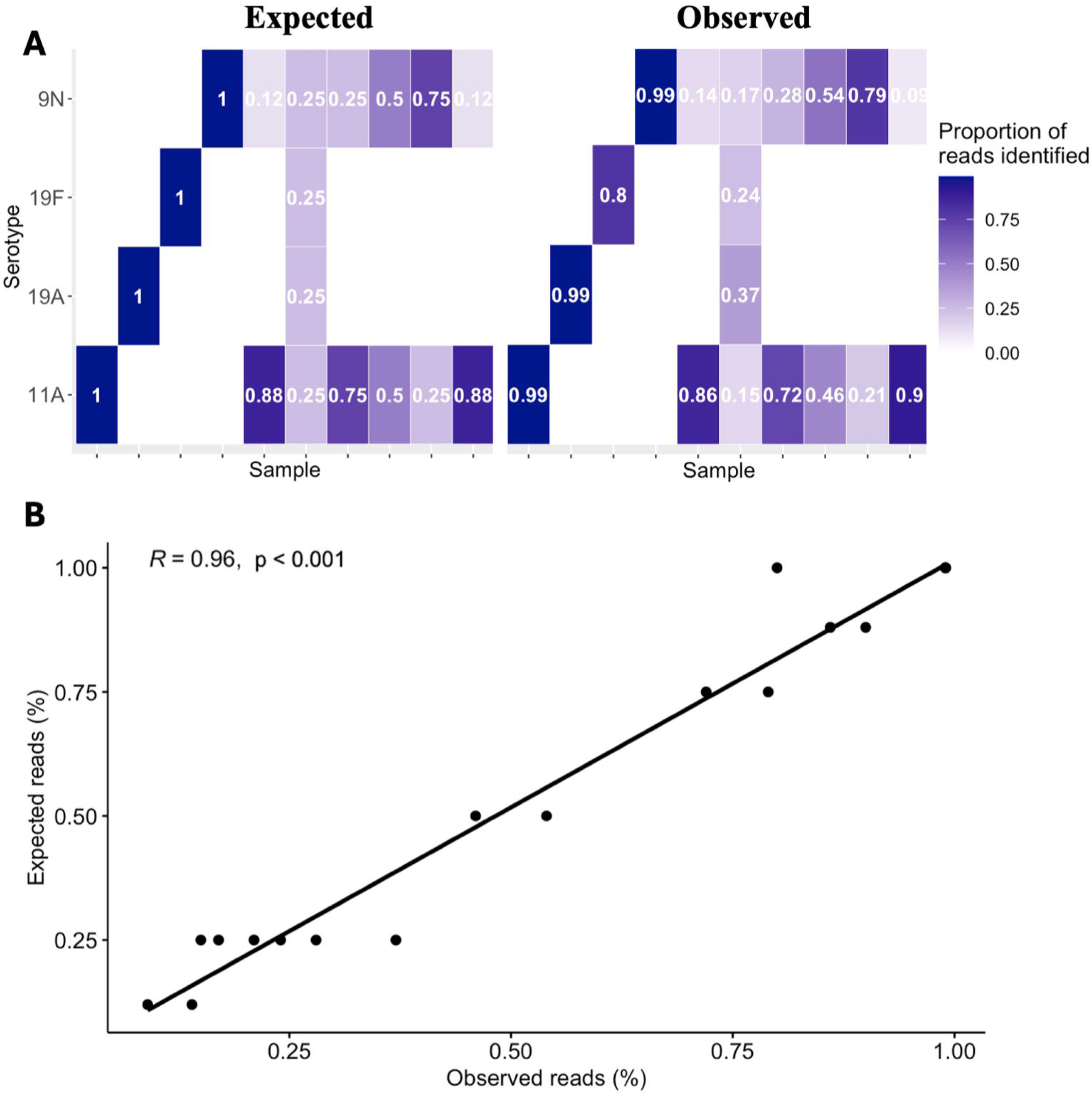
A validation experiment to detect different serotypes in mock samples, made of pure isolate artificial mixes of known serotypes. Serotype specific reads were predicted *in silico*. Both the correct serotype and relative abundance of each could be successfully determined using tNGS.

### 7.3 Target sequence recovery from pleural fluid samples

Five *S.pneumoniae* positive samples failed extraction and consequently sequencing, as very few total pathogen reads were obtained (median=5.3×10^3^, IQR=222-7915). All these samples had Ct values over 32 and Qubit values after library preparation ranged between 0.2-8.5 (median = 4.4) with no detectable gene enrichment and could be an indication of failed library pooling. Due to insufficient amount of residual pleural fluid, we were unable to repeat the extraction. Consequently, these samples were removed from the analysis of tNGS performance. A total of 39 pleural fluid samples were used to validate culture-independent serotyping using tNGS.

The enrichment of *S.pneumoniae* sequences was achieved in PCR positive fluids (n=39) with deduplicated reads mapping to enriched rMLST, and sequetyping *cpsB, ply, lytA* and *SP2020* targets (median deduplicated reads= 5.5×10^2^, IQR=38-2830). Targeted *S.pneumoniae* reads in these samples accounted for over 9.5% of the total bacterial read count, with minimal reads detected for other bacteria (median reads= 3). Target reads aligned with the *cps* sequetyping region in all *S.pneumoniae* positive samples (n=39, median reads = 1.6×10^2^, IQR = 2.5-2046). Deduplicated reads also mapped to three *S.pneumoniae*-specific genes in all but two specimens, *ply* gene (n=37, median = 196, IQR = 1-3448), *lytA* (n=39, median = 143, IQR = 2.5-2108), and *SP2020* (n=38, median = 77, IQR = 1.5-1113). In one sample, the *ply* gene was not detected, and in the other, the *ply* and *SP2020* genes were not detected. Neither of these samples could be serotyped and *S.pneumoniae* PCR cycle threshold (Ct) values were over 30 in both samples.

Deduplicated reads were not enriched for *S.pneumoniae* in fluids which were either negative by PCR or positive for other bacteria. The total number of raw pathogen reads were highest in the *S.pneumoniae* PCR positive samples (median = 5×10^5^, IQR =8535-554228), that possessed genes enriched by the Castanet v2 probe panel. This was followed by other organism positive samples (median = 2.1×10^5^, IQR =27424-35232), with the Castanet probes able to detect the cultured organism in each specimen (total pathogen reads for *Streptococcus vestibularis =* 9.3×10^5^, *Streptococcus intermedius =* 2.7×10^4^, *Corynebacterium striatum =* 2.6×10^4^, *Listeria monocytogenes =* 3.5×10^4^ *Staphylococcus aureus =* 3×10^4^). As expected, fewer deduplicated microbial reads were present in the negative pleural fluid samples (median = 1.9×10^4^, IQR =14955-21103) (Figure 3A). Samples with lower input *S.pneumoniae* DNA load, indicated by the higher Ct values, demonstrated lower numbers of total *S.pneumoniae* deduplicated reads. There was a significant association between the Ct value of input DNA and the success of target gene enrichment, assessed by the total number of deduplicated reads (p <0.001) (Figure 3B).

**Figure 3.**
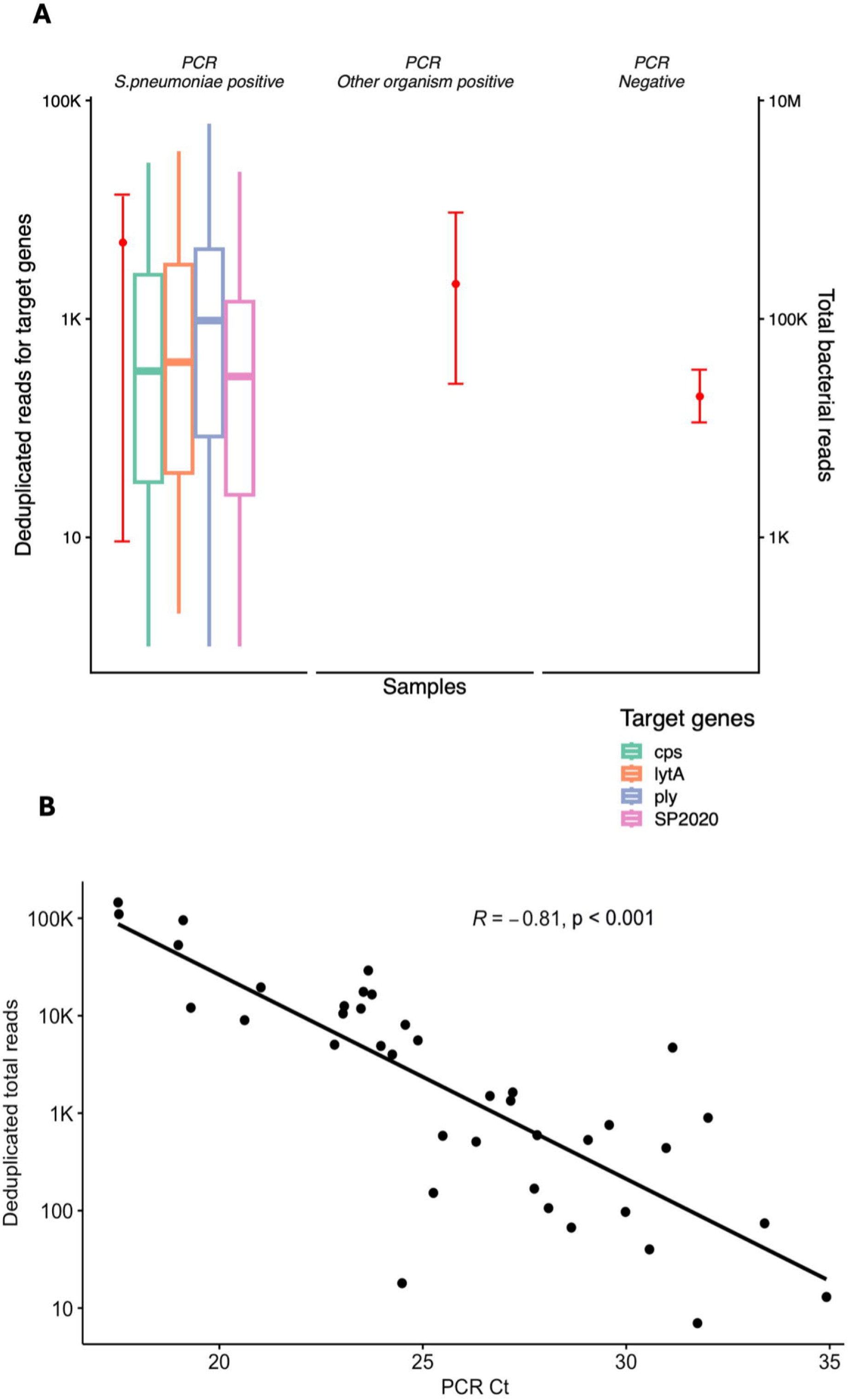
There was successful enrichment of *S.pneumoniae* genes and these were specific to *S.pneumoniae* PCR positive samples only. There was successful sequencing in each category of pleural fluid samples, with few total reads in the pleural fluid negative cases. Note the change in axis for gene specific deduplicated reads on the left and total reads on the right of the plot, and the use of a log scale. The points represent the mean total bacterial read counts with bars representing the minium and maximum total read count for each category. Three outliers with greater than 2 million total bacterial reads that were PCR *S.pneumoniae* positive were not included in the figure (A). There is a correlation between high PCR cycle threshold (Ct) values, indicating low input DNA, and enrichment success (B).

### 7.4 Culture-independent determination of pneumococcal serotype by tNGS

Sufficient sequencing read coverage and depth was achieved in 95% (37/39) of *S.pneumoniae* PCR positive pleural fluids. The median depth for the sequetyping region was 949 (IQR=109-6584, p=0.003) with a median sequetyping region coverage of 100 (IQR=100-100, p <0.001). Sequetyping reconstruction indicated a clear dominance of a single serotype in each known positive sample, with strongly supportive TPM values (median=998,122, IQR=993003-999109, p<0.001) and reference read coverage and depth (Figure 4).

**Figure 4.**
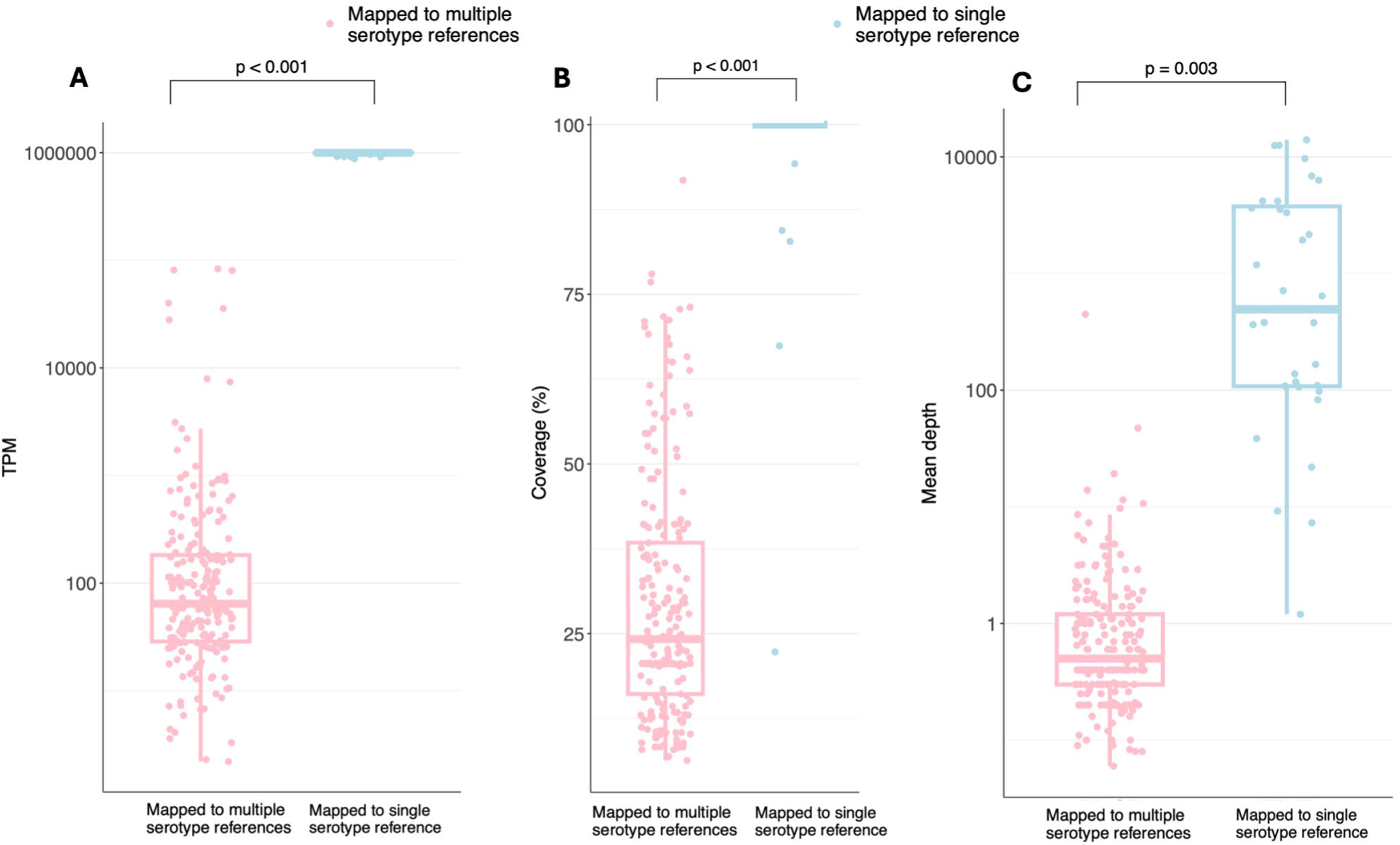
Sequetyping analysis demonstrating the dominance of reads mapping to a single serotype reference for each specimen, compared to remaining reads that mapped to multiple different serotype references. Three bioinformatic parameters were used to determine the serotype within each specimen (A) transcript per million (TPM), (B) percent reference coverage and (C) mean depth. Four outlier results, with greater than 20,000 mean depth, are not included in the figure. Note the change in axis between the plots and TPM and mean depth use a log scale.

tNGS provided *S. pneumoniae* serotyping results for 37 pleural fluids in our cohort and identified 16 different serotypes (Figure 5B). The most common serotype from this selection of unculturable samples, was serotype 3 (12/37 or 32%). In contrast, only 56% of these samples were previously serotyped with standard PCR-based sequetyping (p <0.001) (Figure 5A). Two samples could not be confidently serotyped either by tNGS or standard sequetyping. All serotyping results were concordant with previous molecular results, except for two specimens. One result was identified as 33F by molecular methods, but 33B by tNGS. The other was initially identified as serotype 4 but was resolved to be serotype 3 in this study.

**Figure 5.**
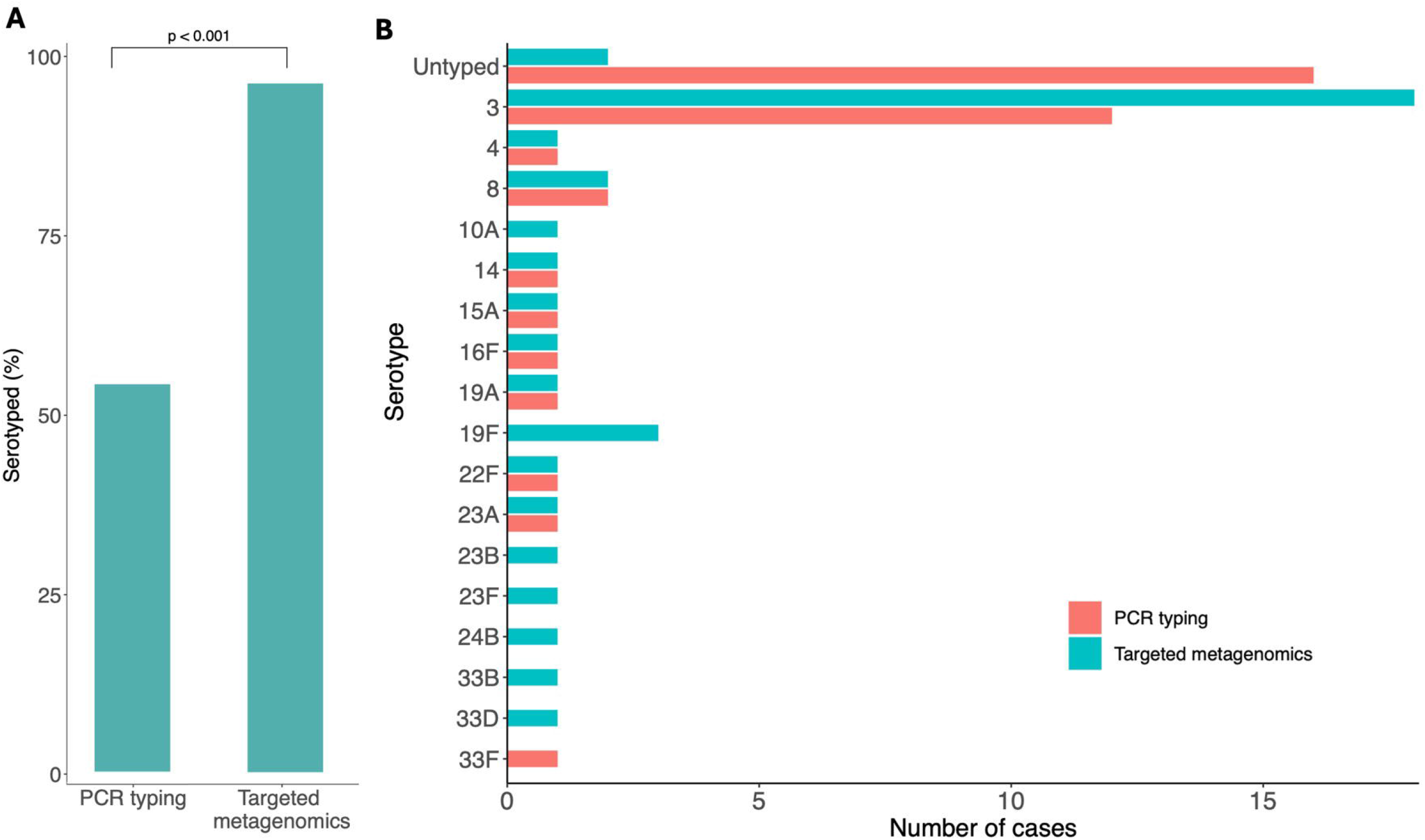
From the study cohort, 56% of cases could be serotyped from molecular methods, compared to tNGS that could serotype 95% (A). tNGS was able to identify a wide range of different serotypes and could distinguish more serotypes compared to molecular methods (B).

## 8. Discussion

This study has designed and validated a highly sensitive targeted metagenomics approach for serotyping *S.pneumoniae* directly from pleural fluid. This method can accurately determine *S.pneumoniae* serotype, and the relative abundance of multiple serotypes within the same sample (Figure 2). Previously, tNGS has been applied for taxonomic identification of bacteria in pleural fluid samples (32), but here we extend the method’s utility to accurately detect *S.pneumoniae*, and determine pneumococcal serotypes in the context of documented IPD. Pleural infections are often polymicrobial (11, 33), and IPD cases associated with co-infection with distinct serotypes of *S.pneumoniae* have experienced increased mortality (34). Such pneumococcal co-infections appear to be rare, are predominately reported in children, and difficult to detect using current molecular methods due to primer mismatches in the *cpsB* sequetyping region (35, 36). Therefore, our tNGS approach offers a sensitive and robust method to overcome this diagnostic and surveillance challenge.

To the best of our knowledge, this is the first methodology to specifically enrich for the *S.pneumoniae cpsB* gene containing the sequetyping region. Our findings demonstrated 100% specificity and sensitivity for detection of *S.pneumoniae* and produce accurate serotyping data based on the *cpsB* gene sequence. Importantly, there was no enrichment for *S.pneumoniae* in known negative specimens or cross hybridisation from pleural fluids positive for other bacteria (Figure 3A). We demonstrated successful *S.pneumoniae* enrichment in all, but two cases. In these two failed specimens we could not detect reads aligned to *ply* or both *ply* and *SP2020* targets, both of which are core *S.pneumoniae* genes. However, these samples had *S.pneumoniae* PCR Ct values of 34.92 and 31.75 respectively. This observation suggests the potential limit of sensitivity for input pathogen DNA load of the tNGS methodology, with our results confirming limited recovery from specimens requiring greater than 30 cycles for positive *S.pneumoniae* PCR (Figure 3B). Our results reconfirmed previous reports where target coverage and depth thresholds were less likely to be achieved with Ct >30 (22, 37, 38). However, lower sensitivity limits, up to Ct of 38, have been reported when the tNGS method was applied for respiratory pathogen detection without characterisation (39).

Our tNGS serotyping approach has been able to successfully serotype 95% (37/39) of samples compared to 56% (22/39) obtained from molecular methods (Figure 5A). There was concordance with serotyping results between the two serotyping techniques, except for two samples. This included a 33F as identified by molecular methods but identified as 33B by tNGS. These two serotypes belong to the same serogroup and hence are genomically more similar (40, 41), possibly explaining the discordant results. A sample was identified as serotype 4 was identified by molecular approaches but was identified as serotype 3 in tNGS. Lack of pleural fluid clinical sample, meant molecular or tNGS methods could not be repeated to confirm this result. It is uncertain why there were discordant results, as there was high mean depth, TPM and 100% coverage.

We further examined potential read misalignments which can occur when reads are mapped to the incorrect sequetyping reference. When reference sequences are genomically similar, pseudo aligners like Kallisto have greater difficulty distinguishing them (42), and closely related or genomically similar sequences, are more difficult to correctly assign reads to (28).

Our findings indicated a clear competitive mapping preference that distinguished the causal serotype in our sample cohort suggesting minimal misalignment in the pleural fluid cases sequenced in this study (Figure 4C). Furthermore, reads mapping to the correct sequetyping region had almost full coverage of the *cpsB* gene (Figure 4B). Mean depth, however, did not have a significant correlation with degree of read misalignment (Figure 4A). Although mixed pneumococcal serotype infections were not documented in our pleural fluid cohort, our testing of synthetic samples with mixed pneumococcal serotypes suggested that the tNGS methodology can accurately distinguish mixed infections and indicate relative abundance of mixed serotypes, making this methodology potentially useful in pneumococcal carriage studies (43, 44).

In the past, PCR assays have had trouble distinguishing between serotypes from the same serogroup such as 7F and 7A (45), which is problematic as 7F is included within the PCV13 while 7A is not. tNGS, however was able to distinguish 16 different serotypes including those within the same serogroup (Figure 5B).

The most common serotype found in pleural samples in our cohort was serotype 3, which is consistent with the distribution of serotypes seen in NSW affecting children under the age of 5 (Figure 1A).Other studies have reported the increase in serotype 3 causing complicated pneumonia in this age group (46). Since the introduction of PCV13, there has been a reported increase in serotype 3 causing IPD with pleural empyema (47). Our findings also suggested that sensitivity of pleural fluid culture might be lower for serotype 3 infections, and as such prevalence in the pleural fluid space is likely to be under reported if serotyping results are based on culture methods alone (48). The use of tNGS to serotype within this clinically important sample type, that causes severe illness especially in children under the age of 5, may provide a higher resolution for testing and surveillance.

Currently pneumococcal serotypes causing empyema cannot be identified in a third of cases as this clinical sample type is difficult to culture. This study sought to fill this surveillance gap, by developing a tNGS method capable of distinguishing serotypes independently of the culture.Technical expertise and access to laboratory infrastructure are necessary for use of tNGS within a routine clinical setting. Though tNGS can detect target DNA at much lower concentrations compared to other molecular methods, there is still a limit of detection and characterisation. When pathogen load is reduced, target-enrichment methods may not be able to capture the gene of interest with sufficient coverage and depth.

In conclusion, our findings validated the use of capture-based enrichment for culture independent pneumococcal serotyping. The proposed tNGS methodology significantly improves the ability to directly identify and serotype IPD-associated serotypes of *S. pneumoniae* in difficult-to-culture pleural fluids. It can enhance resolution of laboratory surveillance of IPD as well as our understanding of pneumococcal vaccine effectiveness.

To understand the dynamics of empyema caused by *S.pneumoniae* further studies with greater sample size are required to reveal the serotype distribution in pleural fluid. Though pleural fluid is the only clinical sample type discussed here this culture-independent serotyping method has the potential to be applied to a broader range of sample types associated with IPD.

## Data Availability

All data produced in present study is contained in the manuscript

https://www.ebi.ac.uk/ena/browser/view/PRJEB111154

## 10. Author statements

### 10.1 Author contributions

Conceptulisation performed by S.A.M.S, R.J.R, V.S. Data curation contributed by S.A.M.S, S.O. Formal analysis performed by S.A.M.S. Funding acquisition from V.S and R.J.R. Investigation by S.A.M.S, K.KG.T, S.O. Methodology contributed by S.A.M.S, M.P, T.G. Project administration performed by S.A.M.S, R.J.R, V.S. Software contributed by M.P, T.G. Supervision from R.J.R and V.S. Validation from R.J.R and V.S. Visualization performed by S.A.M.S. Writing – original draft performed by S.A.M.S, R.J.R and V.S. Writing – review and editing performed by all authors

### 10.4 Ethical approval

Clinical samples and metadata were collected by the PRL at the NSW Health Pathology-Institute of Clinical Pathology and Medical Research under the Western Sydney Local Health District Human Research Ethics and Governance Committee (Project identifier: 2019/PID14240).

