## Supplementary data for "Culture-independent identification and serotyping of *Streptococcus pneumoniae* by targeted metagenomics in pleural fluid samples"

**Supplementary data: Comprehensive *cps* reference sequences with GenBank accession numbers**

| GenBank accession  numbers | Serotypes |
| --- | --- |
| CR931632 | 1 |
| CP000920 | 1 |
| CP001845 | 1 |
| FQ312030 | 1 |
| FQ312039 | 1 |
| FQ312042 | 1 |
| JN642309 | 1 |
| JN660116 | 1 |
| JN660117 | 1 |
| JN660118 | 1 |
| JN660119 | 1 |
| JN660120 | 1 |
| JN660121 | 1 |
| JN660122 | 1 |
| Z83335 | 1 |
| CR931633 | 2 |
| AF026471 | 2 |
| CP000410 | 2 |
| JN660123 | 2 |
| CR931634 | 3 |
| HE983624 | 3 |
| FQ312027 | 3 |
| FQ312041 | 3 |
| FQ312043 | 3 |
| FQ312044 | 3 |
| FQ312045 | 3 |
| JN680145 | 3 |
| JQ743514 | 3 |
| JQ743515 | 3 |
| JQ743516 | 3 |
| JQ743517 | 3 |
| JQ743518 | 3 |
| Z47210 | 3 |
| JN660145 | 3 |
| CR931635 | 4 |
| AE005672 | 4 |
| AF316639 | 4 |
| JN660124 | 4 |
| JN660125 | 4 |
| JN664256 | 4 |
| JN680110 | 4 |
| JN680122 | 4 |
| JN680123 | 4 |
| JQ743519 | 4 |
| JQ743520 | 4 |
| JQ743521 | 4 |
| CR931637 | 5 |
| AY336008 | 5 |
| JQ743525 | 5 |
| CP000918 | 5 |
| JN660126 | 5 |
| JQ743522 | 5 |
| JQ743523 | 5 |
| JQ743524 | 5 |
| JF911488 | 6A |
| JF911490 | 6A |
| JF911493 | 6A |
| JF911499 | 6A |
| JF911505 | 6A |
| JQ743526 | 6A |
| JQ743529 | 6A |
| JF911487 | 6A |
| JF911491 | 6A |
| JF911495 | 6A |
| JF911496 | 6A |
| JQ743527 | 6A |
| JQ743528 | 6A |
| AY078347 | 6A |
| JN680147 | 6A/6B? |
| JF911489 | 6A |
| JF911497 | 6A |
| CR931638 | 6A |
| JF911492 | 6A |
| JF911494 | 6A |
| JN660127 | 6A |
| JN680106 | 6A |
| JN680118 | 6A |
| JN680128 | 6A |
| JN680138 | 6A |
| JN680146 | 6A |
| JQ743530 | 6A |
| CR931639 | 6B |
| JN642311 | 6B |
| JN642314 | 6B |
| KC522500 | 6B |
| KC522501 | 6B |
| KC522502 | 6B |
| AF316640 | 6B |
| JF911498 | 6B |
| JF911500 | 6B |
| JF911501 | 6B |
| JF911502 | 6B |
| JF911503 | 6B |
| JN660128 | 6B |
| JN660129 | 6B |
| JN680105 | 6B |
| JN680136 | 6B |
| JN680137 | 6B |
| JN680142 | 6B |
| AF246897 | 6B |
| CP002176 | 6B |
| JF911504 | 6B |
| JF911507 | 6B |
| JN642310 | 6B |
| JN642312 | 6B |
| JN642313 | 6B |
| JN642315 | 6B |
| JN680143 | 6B |
| JN680148 | 6B |
| EF538714 | 6C |
| HM448897 | 6C |
| JF911509 | 6C |
| JF911510 | 6C |
| JF911515 | 6C |
| JN660130 | 6C |
| HM171374 | 6D |
| KM114241 | 6E |
| KM114229 | 6E |
| KM114235 | 6E |
| KC832410 | 6E |
| KC832411 | 6F |
| KC522494 | 6X |
| KC522495 | 6X |
| KC522496 | 6X |
| KC522497 | 6X |
| KC522498 | 6X |
| CR931643 | 7F |
| JQ743531 | 7F |
| JQ743532 | 7F |
| JQ743533 | 7F |
| JQ743534 | 7F |
| JQ743535 | 7F |
| JQ743536 | 7F |
| JN660086 | 7F |
| JN660131 | 7F |
| CR931640 | 7A |
| CR931641 | 7B |
| CR931642 | 7C |
| JN642316 | 7C |
| JN642317 | 7C |
| CR931644 | 8 |
| AF316641 | 8 |
| JN660132 | 8 |
| JN660133 | 8 |
| JN680121 | 8 |
| JN680139 | 8 |
| JN680144 | 8 |
| AJ239004 | 8 |
| CR931645 | 9A |
| CR931646 | 9L |
| CR931647 | 9N |
| JN660134 | 9N |
| JN660135 | 9N |
| JN660146 | 9N |
| AF402095 | 9V |
| JN660136 | 9V |
| JN660141 | 9V |
| JN680107 | 9V |
| JN680109 | 9V |
| JN680111 | 9V |
| JN680112 | 9V |
| JN680119 | 9V |
| JN680127 | 9V |
| JN680141 | 9V |
| JQ743537 | 9V |
| JQ743538 | 9V |
| JQ743539 | 9V |
| JQ743540 | 9V |
| CR931648 | 9V |
| CR931652 | 10F |
| JN660088 | 10F |
| CR931649 | 10A |
| JN642318 | 10A |
| JN642319 | 10A |
| JN660087 | 10A |
| JN680133 | 10A |
| KC688318 | 10A |
| CR931650 | 10B |
| CR931651 | 10C |
| CR931657 | 11F |
| CR931653 | 11A |
| CP002121 | 11A |
| JN642320 | 11A |
| JN660089 | 11A |
| JX102570 | 11A |
| CR931654 | 11B |
| CR931655 | 11C |
| CR931656 | 11D |
| JX102571 | 11D |
| JX102572 | 11 |
| CR931660 | 12F |
| JN660090 | 12F |
| CR931658 | 12A |
| CR931659 | 12B |
| JN660091 | 12B |
| JN680126 | 12B |
| CR931661 | 13 |
| JQ653093 | 13 |
| CR931662 | 14 |
| JN660142 | 14 |
| JN660143 | 14 |
| CP001033 | 14 |
| CP000919 | 14 |
| FQ312029 | 14 |
| JN660147 | 14 |
| JN680130 | 14 |
| JQ743542 | 14 |
| JQ743541 | 14 |
| X85787 | 14 |
| JN680125 | 14 |
| JN660144 | 14 |
| CR931666 | 15F |
| CR931663 | 15A |
| JN660092 | 15A |
| JN680113 | 15A |
| JN680114 | 15A |
| JN680134 | 15A |
| CR931664 | 15B |
| JN660093 | 15B |
| KC688319 | 15B/15C |
| CR931665 | 15C |
| CR931668 | 16F |
| JN660094 | 16F |
| CR931667 | 16A |
| JN642321 | 17F |
| CR931670 | 17F |
| JN660137 | 17F |
| JN660140 | 17F |
| JN660138 | 17F |
| CR931669 | 17A |
| JN660095 | 17A |
| CR931674 | 18F |
| CR931671 | 18A |
| CR931672 | 18B |
| JN660096 | 18B |
| AF316642 | 18C |
| CR931673 | 18C |
| JQ743543 | 18C |
| JQ743544 | 18C |
| JQ743545 | 18C |
| JQ743546 | 18C |
| JQ743547 | 18C |
| JQ743548 | 18C |
| CR931678 | 19F |
| AF030367 | 19F |
| JF911522 | 19F |
| JF911523 | 19F |
| JF911524 | 19F |
| JF911529 | 19F |
| JF911530 | 19F |
| JN642323 | 19F |
| JN642324 | 19F |
| JN642325 | 19F |
| JN664260 | 19F |
| JN680129 | 19F |
| JF911525 | 19F |
| AF030368 | 19F |
| AF030369 | 19F |
| AF030370 | 19F |
| AF030372 | 19F |
| JF911526 | 19F |
| JF911527 | 19F |
| JF911531 | 19F |
| U09239 | 19F |
| CP001015 | 19F |
| JF911528 | 19F |
| JN664259 | 19F |
| JQ743553 | 19F |
| AF030371 | 19F |
| CP000921 | 19F |
| CP003357 | 19F |
| CP006844 | 19F |
| JQ743552 | 19F |
| HG799504 | 19A |
| CR931675 | 19A |
| AF094575 | 19A |
| JF911512 | 19A |
| CP001993 | 19A |
| HG799488 | 19A |
| HG799505 | 19A |
| JF911511 | 19A |
| JF911514 | 19A |
| JF911516 | 19A |
| JF911517 | 19A |
| JF911519 | 19A |
| JF911520 | 19A |
| JN664258 | 19A |
| JN680108 | 19A |
| JN680120 | 19A |
| JN680135 | 19A |
| JQ743549 | 19A |
| JQ743550 | 19A |
| JQ743551 | 19A |
| JF911513 | 19A |
| JF911518 | 19A |
| JF911521 | 19A |
| CP000936 | 19A |
| JN664257 | 19A |
| JN642322 | 19A |
| CR931676 | 19B |
| CR931677 | 19C |
| CR931679 | 20 |
| JN660097 | 20 |
| JQ653094 | 20 |
| CR931680 | 21 |
| JN642326 | 21 |
| JN680140 | 21 |
| JN680117 | 21B |
| JN680131 | 21B |
| JQ009436 | 21 |
| CR931682 | 22F |
| JN660099 | 22F |
| CR931681 | 22A |
| JN660098 | 22A |
| CR931685 | 23F |
| AF030373 | 23F |
| AF030374 | 23F |
| AF057294 | 23F |
| FM211187 | 23F |
| JN664261 | 23F |
| JN680115 | 23F |
| JN680116 | 23F |
| JN680124 | 23F |
| JN680132 | 23F |
| JQ743554 | 23F |
| JQ743555 | 23F |
| JQ743556 | 23F |
| JQ743557 | 23F |
| JQ743558 | 23F |
| CR931683 | 23A |
| CR931684 | 23B |
| JN642327 | 23B |
| JN660100 | 23B |
| CR931688 | 24F |
| JN660102 | 24F |
| CR931686 | 24A |
| CR931687 | 24B |
| JN660101 | 24B |
| CR931690 | 25F |
| CR931689 | 25A |
| CR931691 | 27 |
| JN660139 | 27 |
| CR931692 | 28A |
| JN660103 | 28A |
| CR931693 | 28F |
| JN660104 | 28F |
| CR931694 | 29 |
| **JN660105** | 29 |
| CR931695 | 31 |
| JN660106 | 31 |
| CR931697 | 32F |
| CR931696 | 32A |
| CR931702 | 33F |
| AJ006986 | 33F |
| JN660111 | 33F |
| JN660107 | 33A |
| CR931698 | 33A |
| CR931699 | 33B |
| JN660108 | 33B |
| CR931700 | 33C |
| JN660109 | 33C |
| HE651321 | 33C/33B |
| CR931701 | 33D |
| JN660110 | 33D |
| CR931703 | 34 |
| JN660112 | 34 |
| CR931707 | 35F |
| CR931704 | 35A |
| JN660113 | 35A |
| JN642328 | 35A |
| CR931705 | 35B |
| JN660114 | 35B |
| CR931706 | 35C |
| CR931708 | 36 |
| JN660115 | 36 |
| AJ131984 | 37 |
| CR931709 | 37 |
| CR931710 | 38 |
| CR931711 | 39 |
| CR931712 | 40 |
| CR931714 | 41F |
| CR931713 | 41A |
| CR931715 | 42 |
| CR931716 | 43 |
| CR931717 | 44 |
| CR931718 | 45 |
| CR931719 | 46 |
| CR931721 | 47F |
| CR931720 | 47A |
| CR931722 | 48 |
| MW732693 | *Oralis* |
| CP034442 | *mitis* |
| OY769914 | *Oralis* |
| CP065707 | *oralis* |
| CP065706 | *oralis* |
| CP046523 | *oralis* |
| LR594040 | *australis* |
| MW732691 | *mitis* |
| MW732690 | *mitis* |
| MZ857152 | *mitis* |
| MZ857150 | *mitis* |
| MW732692 | *mitis* |
| MZ857149 | *mitis* |
| MZ857154 | *mitis* |
